# Using the OHDSI network to develop and externally validate a patient-level prediction model for Heart Failure in Type II Diabetes Mellitus

**DOI:** 10.1101/2021.04.06.21254966

**Authors:** Ross D. Williams, Jenna M. Reps, Jan A Kors, Patrick B Ryan, Ewout Steyerberg, Katia M. Verhamme, Peter R. Rijnbeek

## Abstract

**Introduction:** Heart Failure (HF) and Type 2 Diabetes Mellitus (T2DM) frequently coexist and exacerbate symptoms of each other. Treatments are available for T2DM that also provide beneficial treatment effects for HF. Guidelines recommend that patients with HF should be given Sodium-glucose co-transporter-2 inhibitors in preference to other second-line treatments for T2DM. Increasing personalization of treatment means that patients who have or are at risk of HF receive a customised treatment. We aimed to develop and externally validate prediction models to predict the 1-year risk of incident HF in T2DM patients starting second-line treatment.

**Methods:** We analysed a federated network of electronic medical records and administrative claims data from five databases (CCAE, MDCD, MDCR, Optum Clinformatics and Optum EHR) in the United States. We used each database to develop a model to predict 1-year risk of incident HF in patients initialising a second pharmaceutical intervention, following initial treatment with metformin for T2DM. We then performed internal validation for each model as well as external validation using the other databases.

**Results:** A total of 403,187 patients were included in the study. We developed 5 models with discrimination ranging from 0.72 to 0.80 at external validation in the other databases. Consistent high performance was noted for models developed in CCAE, Optum Clinformatics and Optum EHR with AUCs ranging from 0.74 to 0.81. For these models, calibration was acceptable.

**Conclusion:** Three high-performing prediction models were developed for this problem. The CCAE developed model was selected for recommendation as it achieved the same discrimination and better calibration performance than the Optum Clinformatics and Optum EHR models, whilst selecting fewer covariates and as such was selected as the best developed model. The models could be useful in stratifying patient treatment, planning healthcare utilization and reducing cost by aiding in increasing preparedness of healthcare providers.

## Introduction

Type 2 Diabetes Mellitus (T2DM) and Heart Failure (HF) are two major and growing contributors to the global burden of disease both in developed and developing countries. Globally an estimated 462 million people are affected by T2DM (1) and 64 million by HF(2). The incidence of T2DM is increasing, as demonstrated by a study in the UK (3) where the prevalence of T2DM more than doubled from 2.39% (95% CI 2.37 to 2.41) in 2000 to 5.32% (95% CI 5.30 to 5.34) in 2013. This has a high impact on the National Health Service budget in the UK. The situation is similar in the US with an estimated 30.3 million people of all ages, i.e. 9.4% of the US population, having T2DM in 2015 (4). The percentage of adults with T2DM increased with age, reaching a high of 25% among those aged 65 years or older.

In Europe, a reported 1-2% of total health-care expenditure is being used for the management of HF (5). The risk of HF also increases sharply with age. Similar prevalence changes were reported in the US (6). There has been a reduction in HF mortality, however the total burden of disease continues to grow and is expected to increase as the global population ages (7).

T2DM and HF frequently co-exist and research into their bi-directional relationship receives a lot of attention (8, 9). Multiple studies have shown that HF is a major contributor to cardiovascular morbidity and mortality in patients with T2DM, but also conversely those with HF are at higher risk of developing T2DM (10). There is an increased recognition that diabetic patients develop HF independent of the presence of coronary artery disease or its associated risk factors (11). Patients with T2DM are much more likely to develop congestive HF than patients without (9). HF and peripheral arterial disease are the most common initial manifestations of cardiovascular disease in patients with T2DM (12). Patients with T2DM are also more likely to be hospitalized with HF (13, 14).

As described in detail in a literature review, the pathophysiological connection between both diseases and their frequent adverse interactions should impact treatment choice (15). The 2017 (16) and 2018 (17) American Diabetes Association guidelines included recommendations that at initial diabetes diagnosis, the patient should receive a treatment with metformin and lifestyle interventions. If the patient’s HbA1C level remains above 9% after three months then a further drug should be added and this should be assessed based upon the risk profiles of the drugs which best suit the patient. In the 2019 guidelines (18) the advice is updated to stratify patients based upon established HF, atherosclerotic cardiovascular disease and chronic kidney disease. Specifically, the guidelines state that thiazolidinediones (TZD) should be avoided in patients with heart failure and that in patients at high risk of heart failure Sodium-glucose co-transporter-2 inhibitors (SGLT2i) are preferred. The guidelines also recommend different medications for patients who have or are at risk of other diseases. For example, diabetes patients with atherosclerotic cardiovascular disease and low estimated glomerular filtration rate are recommended to be treated with GLP-1 RA. The guidelines appear to be trending towards a more personalized treatment strategy and as such there is an opportunity to use risk prediction to further personalize treatment in the intermediate steps before treatment with insulin use is needed. In a review of prediction studies (19), multiple prediction models for incident HF in T2DM were analyzed, however many of these were set at a later stage of diabetes disease progression or they contained information that was not well captured in the available claims healthcare databases, for example alcohol usage and education level.

The aim of this study is to develop and externally validate a prediction model for predicting 1-year risk of incident HF in patients with T2DM who initiate a second pharmaceutical intervention for control of their diabetes. These prediction models might aid in the decision making of patient and clinician when changing the pharmaceutical therapy for diabetes.

## Methods

This retrospective cohort study used observational healthcare databases from the United States. We developed a model to predict the 1-year risk of HF in T2DM patients who start secondary treatment for T2DM (after Metformin). We developed five models using data from five different US data sources.

### Data Sources

The analyses were performed across a network of five observational healthcare databases. All databases contained either claims or EHR data from the US and have been transformed into the OMOP Common Data Model (OMOP CDM), version 5 (20). Table 1 describes the databases that are included in this study. The complete specification for the OMOP CDM, version 5 is available at: https://ohdsi.github.io/CommonDataModel/cdm531.html

**Table 1.** Database characteristics

| Database | Acronym | Country | Data type | Time period | Database size<br>(million patients) |
| --- | --- | --- | --- | --- | --- |
| Optum® de-identified<br>Electronic Health<br>Record Dataset | Optum EHR | US | EHR | 2006-2018 | 87 |
| IBM MarketScan®<br>Commercial Database | CCAE | US | Claims | 2000-2018 | 155 |
| IBM MarketScan®<br>Multi-State Medicaid<br>Database | MDCD | US | Claims | 2006-2017 | 30 |
| IBM MarketScan®<br>Medicare<br>Supplemental<br>Database | MDCR | US | Claims | 2000-2018 | 10 |
| Optum® De-Identified<br>Clinformatics® Data<br>Mart Database | Optum<br>Clinformatics | US | Claims | 2000-2018 | 98 |

### IRB approval

The use of IBM and Optum databases were reviewed by the New England Institutional Review Board (IRB) and were determined to be exempt from broad IRB approval.

### Cohort definitions

#### Target Cohort

The target population consisted T2DM patients who are treated with metformin who are become new adult users of one of Sulfonylureas, Thiazolidinediones, Dipeptidyl peptidase-4 inhibitors, Glucagon-like peptide-1 receptor agonists, or SGLT2is. The index date is the first prescription of one of these secondary treatments. We required all subjects to have a T2DM diagnosis, which was based upon the presence of a disease code and use of Metformin prior to the index date. Patients with HF or patients treated with insulin on or prior to the index date were excluded from the analysis. Patients were required to have been enrolled for at least 365 days before cohort entry.

#### Outcome definitions

The outcome was defined using the presence of a diagnosis code of HF in occurring for the first time in the patient’s history, between the index date and the end of the time at risk.

The cohort definitions are available at: https://github.com/ohdsi-studies/PredictingHFinT2DM/tree/main/validation/inst/cohorts

The study used a time at risk from 1 to 365 days post index date.

The study period contained data from 2000-2018. The exact period varies between the databases and is available in Table 1.

## Analysis Methods

Model development, per database, followed the framework for the creation and validation of patient-level prediction (PLP) models presented in Reps et al. (21). We used a ‘train-test split’ method to perform internal validation. In each target population cohort, a random sample of 75% of the patients (‘training sample’) was used to develop the prediction model and the remaining 25% of the patients (‘test sample’) was used to internally validate the prediction models developed.

We used regularized logistic regression risk models, also known as least absolute shrinkage and selection operator (LASSO). Regularisation is a process to limit overfitting in model development. This process works by assigning a “cost” to the inclusion of a variable and the variable must contribute more to the model performance than this cost in order to be included. If this condition is not met then the coefficient of the covariate becomes 0, which therefore eliminates the covariate from the model providing an in-built feature selection (22).

### Candidate covariates

We developed a data-driven model using age, sex and binary variables indicating the presence or absence of comorbidity (based on presence of disease codes) any time prior to index (= start of second line treatment) and procedures or drugs that occurred in the year prior to index date. The binary variables were constructed for any condition/procedure/drug that were in the history of any patient in the target cohort. For example, if any patient has a diagnosis of liver failure recorded in their medical records prior to the index date, then we create a candidate binary variable named ‘liver failure any time prior’ that has a value of 1 for patients with a record of liver failure in their history and 0 otherwise. This is done for any condition/procedure/drug. In total, we derived around 39,000 candidate covariates. This included more than 26,000 historical conditions, 13,000 historical treatments, and demographic information.

In addition to this we developed a baseline model that uses only age and sex. This is motivated by a desire to see the added value of a more complex model. The baseline model provides a theoretical minimum performance which can then be compared to the performance of the data-driven model. This provides a theoretical bounding of potential performance.

### External validation

A significant advantage in this study was the availability of multiple datasets. This enabled us to rapidly perform external validation of the prediction models following the framework detailed in Reps et al. (23). To do this we created a model per database and then validated this model both internally and also externally in the other 4 databases. A diagram of this process can be seen in Figure 1. The databases in this study include both claims and EHR, thus the external validation will provide some insight into the generalizability and heterogeneity within the database population and across different care settings.

In total we created 5 unique models each with 4 external validations.

**Figure 1.**
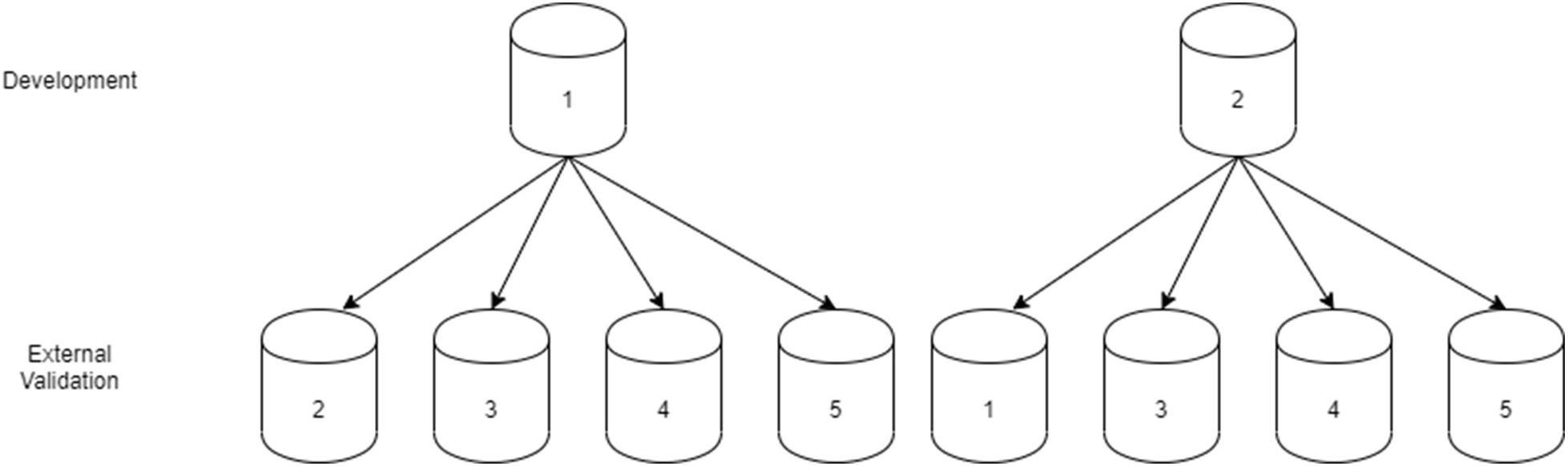
Showing the rotation of databases for model development and external validation. This process was repeated 5 times until a unique model had been developed in each databases and then externally validated 4 times.

### Evaluation Analysis

For performance analysis we consider the area under the receiver operator curve (AUC) as a measure of discrimination. An AUC of 0.5 corresponds to a model randomly assigning risk and an AUC of 1 corresponds to a model that can perfectly rank patients in terms of risk (assigns higher risk to patients who will develop the outcome compared to those who will not). For calibration assessment we used calibration graphs and visually assessed whether the calibration was deemed to be sufficient.

We used the PatientLevelPrediction R-package (version 4.0.1) and R (v4.0.2) to perform all analyses. All development analysis code and cohort definitions are available at: https://github.com/ohdsi-studies/PredictingHFinT2DM

The validation package is available here:

https://github.com/ohdsi-studies/PredictingHFinT2DM/tree/main/validation

## Results

Across all databases we selected 403,187 T2DM patients initiating second-line treatment. Of these, 12,173 developed HF during the one-year follow-up. Next, patient-level prediction of HF was performed. The number of patients and the AUCs are given in Table 2.

**Table 2.** Number of patients and internal validation performance per database

| Database | No. of T2DM patients | No. of HF patients | Incidence (%) | Age in years Mean (SD) | Female (%) | Full model AUC | Age Sex AUC |
| --- | --- | --- | --- | --- | --- | --- | --- |
| ccae | 112,989 | 1,843 | 1.6 | 53 (8) | 46 | 0.78 | 0.64 |
| mdcd | 15,860 | 650 | 4.1 | 50 (12) | 64 | 0.77 | 0.65 |
| mdcr | 22,433 | 1,658 | 7.4 | 73 (6) | 48 | 0.73 | 0.64 |
| Optum Clinformatics | 92,272 | 4,332 | 4.7 | 63 (13) | 48 | 0.80 | 0.69 |
| Optum EHR | 159,633 | 3,690 | 2.3 | 58 (12) | 49 | 0.81 | 0.71 |

**Figure 1.**
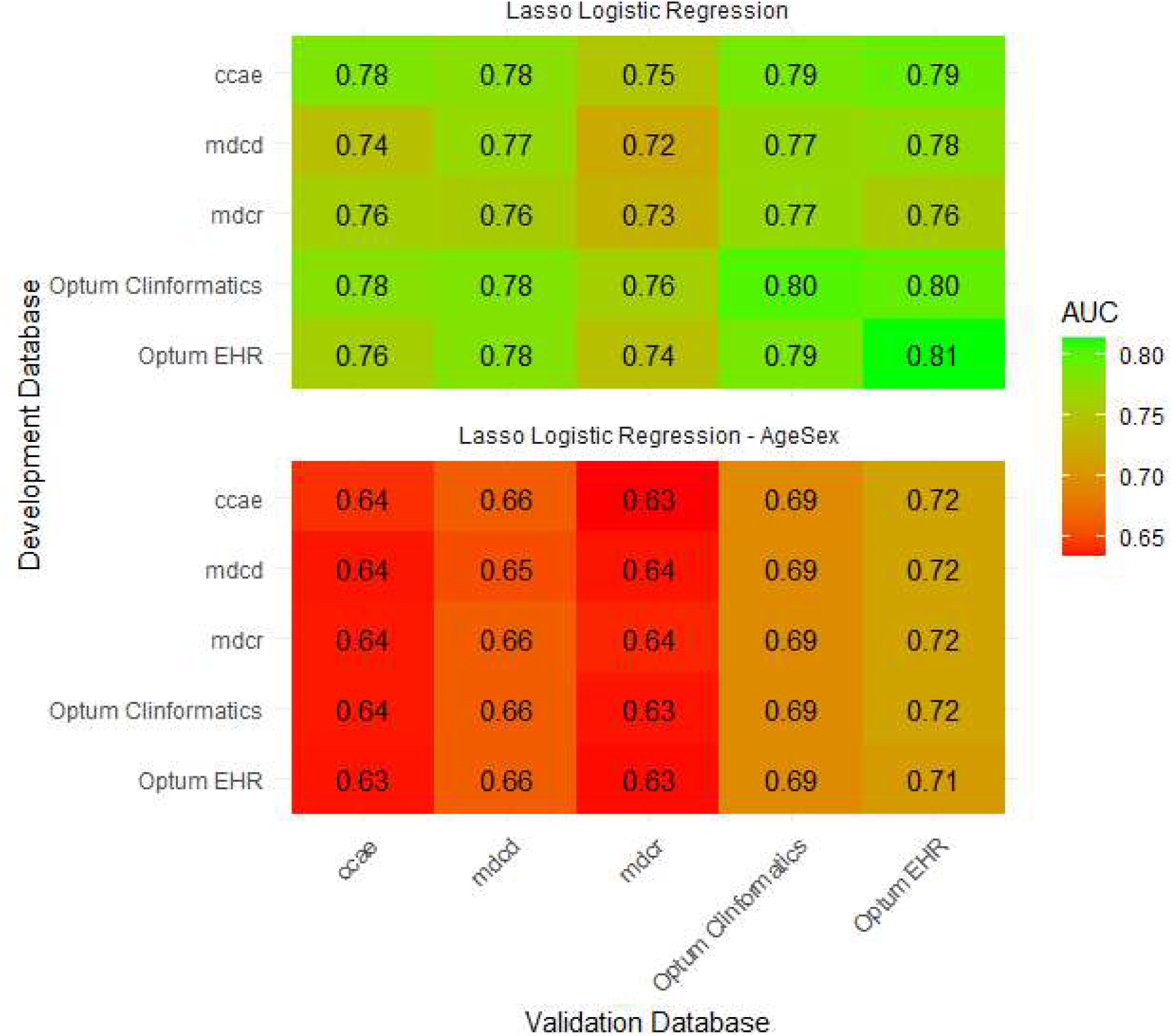
A heatmap of the AUC (95%CI) values across internal and external validations of the developed prediction models. Each row in the figure represents one model, and each column the performance in that database. When the row and column models are the same the result comes from the internal split validation. All other results are from external validation

The AUC results show reasonable performance. The mean AUCs across internal and external validation were 0.78 (CCAE), 0.76 (MDCD), 0.76 MDCR, 0.78 (Optum Clinformatics) and 0.78 (Optum EHR). The best performing models in terms of discrimination were developed in CCAE, Optum ClinFormatics and Optum EHR and appear to be the most consistent across the external validations. As expected, when comparing the baseline model with the full model the performances drop when using only age and sex as predictors.

We assessed the calibration of the three models with the best discrimination (CCAE, Optum Clinformatics and Optum EHR). The calibration results from these 3 models across the external validations are shown in Figure 2. The models generally appear to be well calibrated. These three models had similar discrimination.

**Figure 2.**
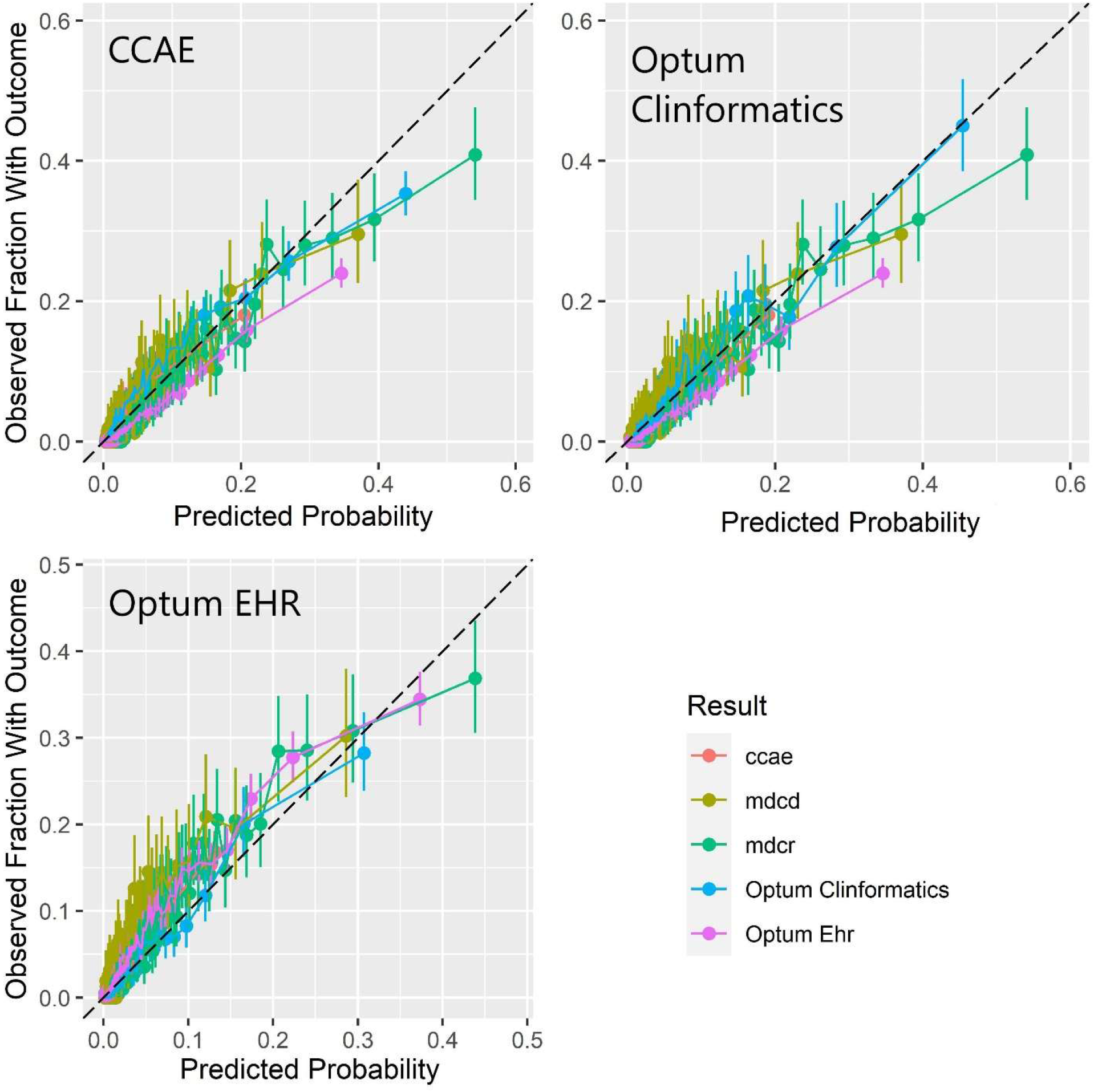
Internal and external calibration of the CCAE, Optum Clinformatics and Optum EHR trained models

The final CCAE model contained 195 covariates, the names and coefficients are available in Appendix 1. All results are available in a study application located at: https://data.ohdsi.org/PredictingHFinT2DM/

## Discussion

This study shows the development of multiple prediction models for HF in T2DM. Of the three highest performing models, the model developed in CCAE had a smaller covariate set (195 for CCAE compared to 413 for Optum Clinformatics and 381 for Optum EHR) and as such was selected for recommendation. The ADA 2019 diabetes guidelines recommend prescribing SGLT2i as second-line treatment to patients at high risk of HF. A well-calibrated model that can predict the risk of HF in patients with T2DM who are about to initiate second-line therapy could therefore be used by clinicians to help identify which patients are likely to benefit from SGLT2i. These patients may then benefit from the protective effects of SGLT2i on HF.

Given the performance of the CCAE model developed in this paper, our model could be used in treatment planning. Indeed, our study demonstrates that this model has good discriminative performance that is consistent across most external validations (AUC internal: 0.78, external 0.75-0.79). There is a minor loss in discrimination for some of the external validations, for example MDCR has the lowest AUC (0.75). This lower performance is in-line with the databases internal validation, and MDCR performs worst across all the external validations suggesting it is a more problematic dataset in which to make predictions. Possible explanations of this are that the underlying case-mix of patients could mean discrimination is harder. For example, patients in this database are generally sicker and as such it could become more difficult to separate them, there is also little to no overlap in ages of patinets between CCAE and MDCR. Another reason could be the lower numbers of patients might mean there is insufficient data to provide a true estimate. The model showed reasonable calibration across internal and external validations with some overestimation of risk for the higher risk patients. The Optum EHR external validation showed a larger miscalibration and could benefit from some recalibration before implementation. Calibration is important when using a model for clinical decision making and this result highlights that our model likely requires recalibration when applied to case-mixes that differ from the development database. To our knowledge this is the only model that is available in open source that can be used for this specific prediction problem.

The baseline age/sex models had only moderate performance across all the validations for all models. Age and sex alone are not sufficient to accurately predict future HF and more complex models are needed.

A major strength of this study is the inclusion of five databases containing large numbers of T2DM patients. This allows for robust external validation of the models.

Limitations include that the model contains 195 variables, which means that it would be difficult to implement this prediction model in the clinic without some automated data input. Another potential limitation is that we did not account for any treatment received after the index date. This might introduce a bias in case HF preventative treatment is initiated in patients at high risk of HF. A related possible bias is the potential misclassification of T2DM or HF. Particularly in the case of HF, if there is misclassification of date of HF diagnosis this could influence the results. The model was also validated in multiple claims but only one EHR database, so it could benefit from further validation in this setting. To assist in this we provide an open software package that can apply our model to any database mapped to the OMOP CDM.

This model selected is in theory the maximum internal performance a logistic regression model could achieve in this setting. This allows for a comparison to be made between this model and a more parsimonious model. This more parsimonious model would be more implementable than the current model in direct clinical practice. The task is to then determine what the acceptable trade-off is between model complexity and performance.

The performance of the selected model suggests that it could be useful in strategic planning. This could occur be either at a treatment facility or health authority level. It will allow these policy makers to assess their T2DM patient population for HF risk and then plan accordingly. This risk-stratified planning could be beneficial both in terms of healthcare outcomes and economical efficiency of treatment.

## Conclusion

Using large regularized regression technique, a good prediction model can be identified. However, there are clinical challenges with implementation of the model in clinical practice to assist patient treatment and management.

## Data Availability

Due to patient privacy concerns no data from the study is available, but upon request access can be discussed. All analysis files and summary statistics form the research will be made available.

